# Assessing long-term pleiotropic effects of potential novel triglyceride-lowering medications using variants identified by Mendelian randomization

**DOI:** 10.1101/2025.03.07.25323588

**Authors:** Elliot Outland, Yi Xin, Alyson L. Dickson, Sergio Mundo, Ran Tao, Xue Zhong, Gul Karakoc, Sevim Serley, Lan Jiang, Nancy J. Cox, Wei-Qi Wei, C. Michael Stein, QiPing Feng

## Abstract

Drugs targeting triglyceride (TG)-associated genes have the potential to improve cardiovascular outcomes. However, we know little regarding the potential additional benefits or deleterious effects of such targeting. As such, in ancestry-specific cohorts of European ancestry (EA) and African ancestry (AA) at Vanderbilt (BioVU), we used PheWAS to test associations between measured TG levels and validated variants previously associated with TG levels in Mendelian randomization studies. We replicated results in All of Us (AoU). In the BioVU EA cohort (n=63,094), 9 of 10 validated SNPs had significant or suggestive associations with lipid and cardiovascular phenotypes, largely consistent with AoU participants of EA (n=97,545). In the BioVU AA cohort (n=12,515) and AoU AA cohort (n=31,710), results were more limited; only 1 of 6 validated SNPs was significantly associated with a lipid or cardiovascular phenotype in either BioVU or AoU, and no significant or suggestive associations were consistent across both cohorts. We detected few secondary effects in either EA or AA BioVU patients, and none were replicated. These results suggest that there may be limited additional benefits, but few deleterious effects, associated with targeting known TG-associated genes. However, these targets may not be as effective for mitigating cardiovascular risk among individuals of AA.

## INTRODUCTION

Elevated triglyceride (TG) levels are associated with an increased risk of coronary heart disease (CHD), the leading cause of death globally.[1,2] In the United States, 31% of adults have triglyceride (TG) levels ≥150 mg/dL (i.e., mildly elevated and above),[2] with studies demonstrating that the risks for CHD increase 14% for men and 37% for women for each mmol (88 mg/dL) rise in TG levels.[3] Indeed, targeting TG can have benefits for reducing CHD risk, even among patients already receiving standard lipid-lowering treatments (e.g., statins, fibrates, omega 3 fatty acids, and niacin).[4–6]

As such, identifying effective therapeutic targets associated with TG levels is an important goal for the reduction of CHD risk; however, successful targeting requires not only a decrease in TG levels, but also an associated secondary effect of improved cardiovascular outcomes. The degree to which and the mechanisms whereby TGs are associated with CHD has been the subject of debate, particularly given the potential for confounding and the interrelated effects of other lipid traits (e.g., low density lipoprotein cholesterol [LDL-C] and apolipoprotein B [ApoB]).[2] One relatively recent methodological approach has offered a means to identify whether TGs are in the causal pathway of CHD more accurately—namely, Mendelian randomization (MR). MR seeks to mirror clinical trials (although it differs in some aspects, including susceptibility to Type 1 selection bias), but rather than random assignment, randomization is achieved through the arbitrary assignment of gene variants passed from parents to offspring.[7] Beneficially, this approach offers the ability to assess associations between exposures and outcomes while limiting unmeasured confounding, since genetic information is fixed at conception.[8–13] Indeed, applying such an approach, researchers have used functional variations in genes with robust effects on TG levels (e.g., *LPL*, *APOA5,* and *APOC3*) to establish that TGs are on the causal pathway of CHD.[12,14–19] These insights have spurred new drug development, targeting specific TG-associated genes for more effective CHD prevention.[20] In particular, current efforts have focused on increasing lipoprotein lipase (LPL) activity; both the *LPL* gene and genes in the LPL pathways (e.g., *ANGPTL3*, *ANGPTL4*, and *APOC3*) have been considered as potential drug targets.[14,21–27]

In addition to establishing these associations, it is also possible to assess other secondary effects of targeting TG-associated genes identified by MR, including adverse effects. During the approval process, clinical trials are designed to test not just the efficacy of new treatments, but also to identify potential side effects (or lack thereof). However, not all phenotypes (effects) can be tested in clinical trials given their relatively limited power and the short periods of follow-up that can obscure long-term effects; these effects typically are only later detected by case reports and retrospective cohort studies of patients that have experienced those effects (e.g., the discovery of cardiovascular risk associated with rofecoxib[28,29]). For example, statins are among the most extensively studied drugs in history and millions of patients take them; however, their increased risk of diabetes was only identified after statins had been on the market for 21 years.[9] Beneficially, combining MR results with PheWAS approaches can facilitate hypothesis generation for potential effects by screening the phenotypic consequences of genetically proxied drug target modulation,[30] offering the advantage of pre-emptively detecting potential long-term effects of such therapies. More specifically, this approach can help with the initial identification of broad on-target effects, even if the primary driver of the causal pathway is another highly-correlated lipid trait. As such, this framework (i.e., MR results combined with PheWAS) presents researchers with the opportunity to generate hypotheses regarding other potential beneficial or deleterious effects of TG-lowering treatments under development.[31] Given that TG-associated genes (particularly *LPL*) are implicated in numerous pathological conditions (e.g., atherosclerosis, diabetes, pancreatitis, and Alzheimer’s disease and related dementias [ADRD]),[32–35] targeting such genes with TG-lowering treatments may have additional effects on these and other conditions. Such approaches are a crucial component of necessary monitoring for potential pleiotropic effects—both beneficial or deleterious) by modelling the real-world deployment of TG-lowering drugs and assessing how known targets may impact patient outcomes within and across populations based in the United States.

Despite this promise, studies of non-CHD effects associated with either TG levels or candidate drugs for TG-lowering have remained limited.[36] In particular, the research to date has focused almost exclusively on patients of European ancestry (EA), predominantly from Finnish and British cohorts;[37–39] this relatively narrow emphasis has potentially hindered our ability to detect effective TG-related targets and predict associated secondary effects, given that lipid profiles can vary significantly by ancestry. Notably, average triglyceride levels are lower in populations of African ancestry (AA) than those of EA,[40] including the lower proportion of individuals with very high TG levels.[40–46] Although genetic studies on inter-individual TG variability have begun to incorporate ancestries beyond those of patients of EA,[46–48] relatively little is known about the relationship between TG-associated genes and secondary effects in individuals of AA. Accordingly, it is important to determine whether associations validated among patients of EA also apply to patients of AA as well as to determine whether there are ancestry-specific secondary effects (either beneficial or deleterious).

As such, to more broadly assess the potential benefits and risks for TG-lowering medications currently in development, we deployed PheWAS to assess the potential pleiotropic effects of MR-identified genetic predictors of TG levels in ancestry-specific cohorts of patients of EA and AA at Vanderbilt University Medical Center [VUMC], with replication among All of Us Research Program [AoU] participants (**Figure 1**). First, we validated associations between 12 genetic variants reported in previous MR studies from 5 genes identified as TG-lowering targets (i.e., *APOA5*, *LPL*, *APOC3*, *ANGPTL3*, and *ANGPTL4*)[37,49] and measured TG levels. Then, among the validated variants, we conducted PheWAS to assess whether these variants were associated with selected prespecified 1) lipid phenotypes, 2) cardiovascular phenotypes, and 3) other phenotypes previously associated with TGs (including diabetes, pancreatitis, and ADRD), as well as 4) unspecified other phenotypes (i.e., without previously reported associations with TGs). Given the complexity of the causal pathway and the derivation of candidate SNPs (from predominantly EA cohorts), we conducted multiple secondary analyses, including additional adjustments for measured LDL-C, genetic variant set-based tests, predicted gene expression testing, interaction assessments among significant results, and interrogation of instrumental variables (IVs) for TGs derived from Global Lipids Genetics Consortium (GLGC)[50–55] cohorts of individuals with AA.

**Figure 1.**
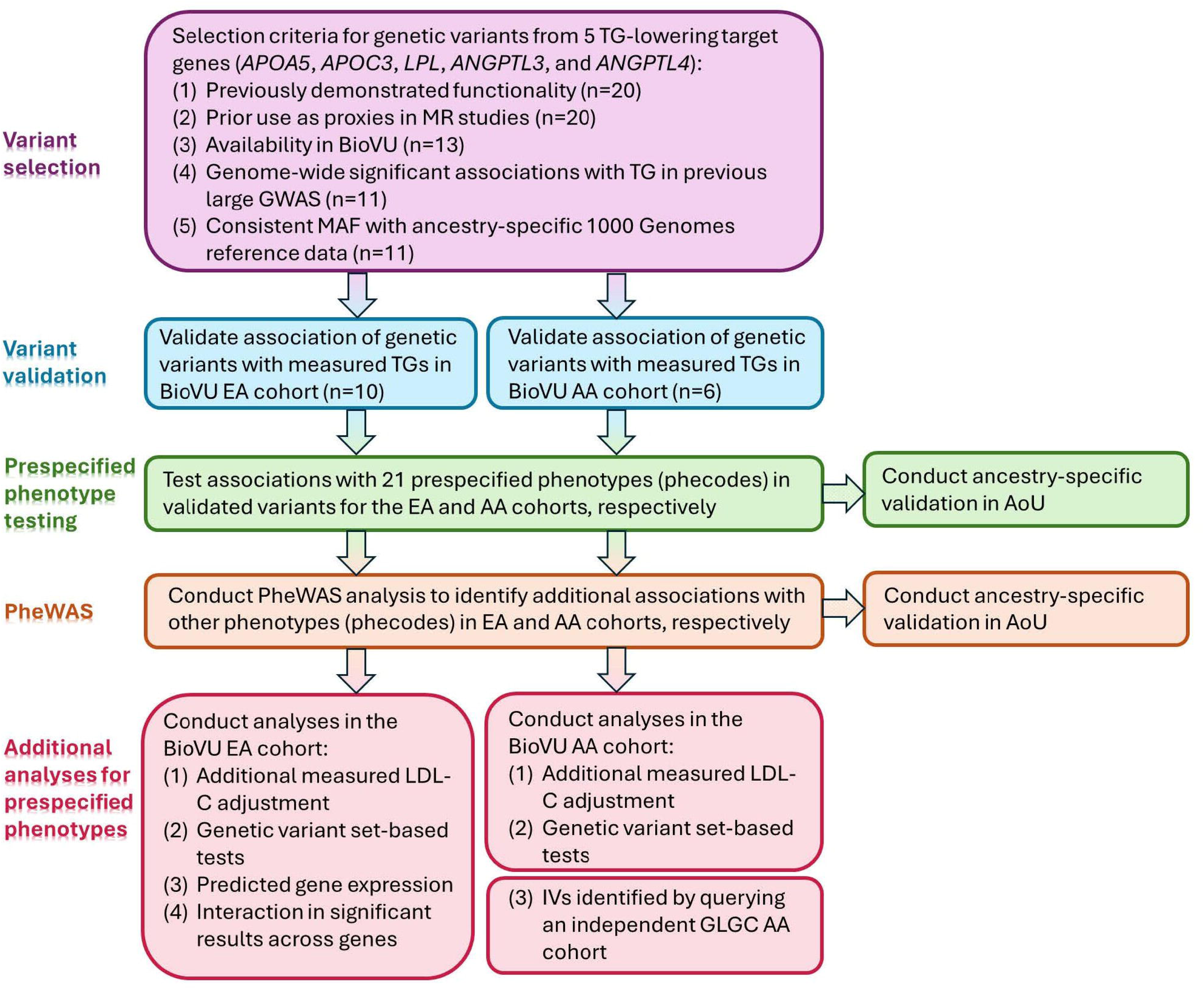
Study design. TG = triglyceride. MR = Mendelian randomization. GWAS = genome-wide association study. MAF = minor allele frequency. EA = European ancestry. AA = African ancestry. PheWAS = phenome-wide association study. BioVU = Vanderbilt University Medical Center biobank. AoU = All of Us Research Program.

## RESULTS

### Cohort characteristics

The EA cohort from BioVU included 63,094 individuals; 35,323 (56.0%) were female, the mean age at the most recent visit was 54.4 ± 21.8, and the mean length of electronic health record (EHR) was 12.0 ± 7.9 years (**Table 1**). The AA cohort from BioVU included 12,515 individuals; 7,737 (61.8%) were female, the mean age was 45.2 ± 20.5, and the mean length of EHR was 11.8 ± 8.7 years (**Table 1**). The adjusted median TG levels for the EA and AA cohorts were 160.9 ± 105.9 mg/dL and 125.5 ± 88.9 mg/dL, respectively. Characteristics for the replication EA (n=97,545) and AA (n=31,710) cohorts in AoU are available in **Supplementary Table 1**.

**Table 1.**
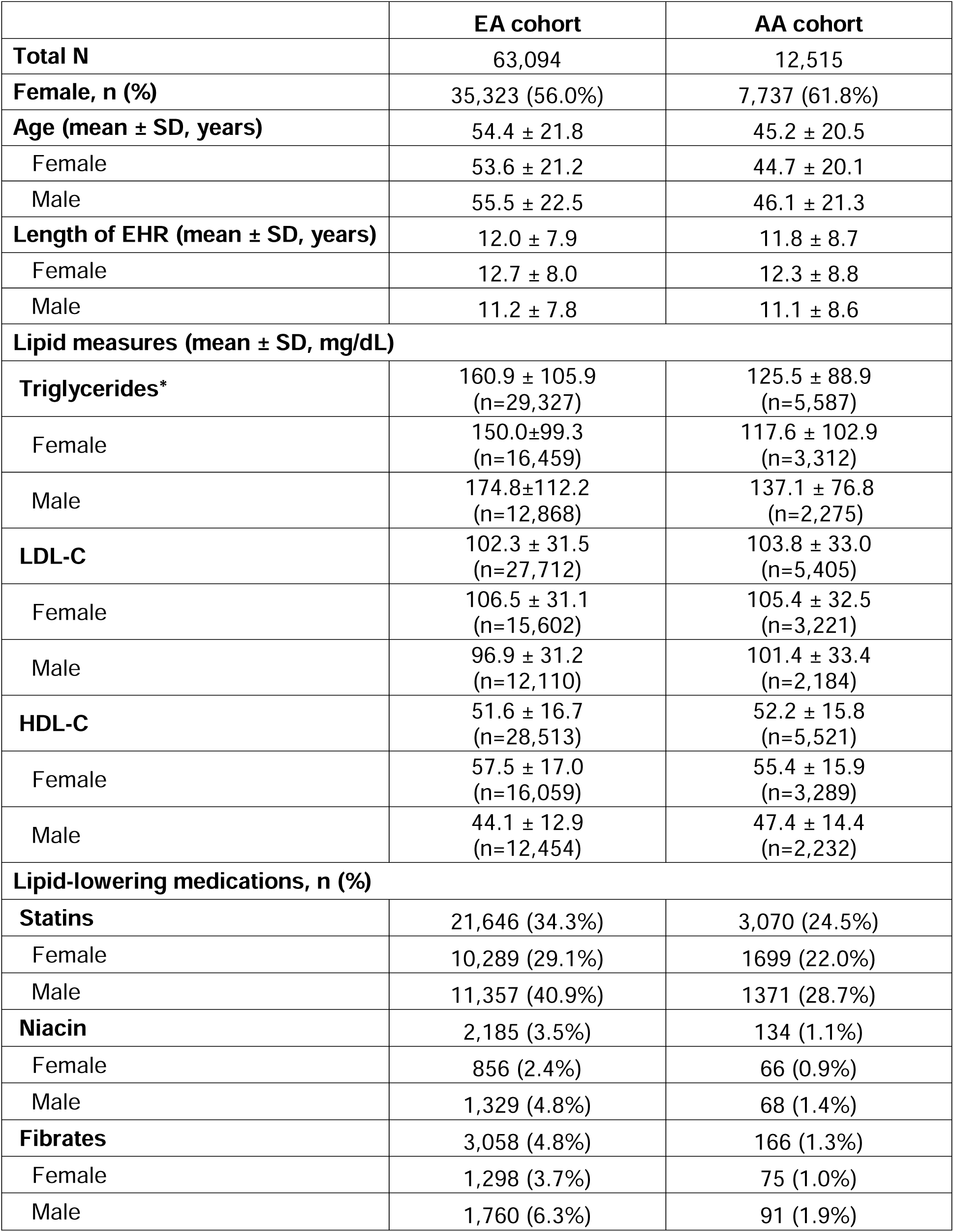
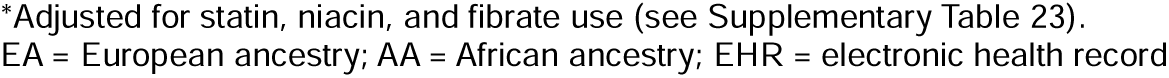
Demographic summary of BioVU cohorts.

|  | **EA cohort** | **AA cohort** |
| --- | --- | --- |
| **Total N** | 63,094 | 12,515 |
| **Female, n (%)** | 35,323 (56.0%) | 7,737 (61.8%) |
| **Age (mean ± SD, years)** | 54.4 ± 21.8 | 45.2 ± 20.5 |
| Female | 53.6 ± 21.2 | 44.7 ± 20.1 |
| Male | 55.5 ± 22.5 | 46.1 ± 21.3 |
| **Length of EHR (mean ± SD, years)** | 12.0 ± 7.9 | 11.8 ± 8.7 |
| Female | 12.7 ± 8.0 | 12.3 ± 8.8 |
| Male | 11.2 ± 7.8 | 11.1 ± 8.6 |
| **Lipid measures (mean ± SD, mg/dL)** |  |  |
| **Triglycerides\*** | 160.9 ± 105.9<br>(n=29,327) | 125.5 ± 88.9<br>(n=5,587) |
| Female | 150.0±99.3<br>(n=16,459) | 117.6 ± 102.9<br>(n=3,312) |
| Male | 174.8±112.2<br>(n=12,868) | 137.1 ± 76.8<br>(n=2,275) |
| **LDL-C** | 102.3 ± 31.5<br>(n=27,712) | 103.8 ± 33.0<br>(n=5,405) |
| Female | 106.5 ± 31.1<br>(n=15,602) | 105.4 ± 32.5<br>(n=3,221) |
| Male | 96.9 ± 31.2<br>(n=12,110) | 101.4 ± 33.4<br>(n=2,184) |
| **HDL-C** | 51.6 ± 16.7<br>(n=28,513) | 52.2 ± 15.8<br>(n=5,521) |
| Female | 57.5 ± 17.0<br>(n=16,059) | 55.4 ± 15.9<br>(n=3,289) |
| Male | 44.1 ± 12.9<br>(n=12,454) | 47.4 ± 14.4<br>(n=2,232) |
| **Lipid-lowering medications, n (%)** |  |  |
| **Statins** | 21,646 (34.3%) | 3,070 (24.5%) |
| Female | 10,289 (29.1%) | 1699 (22.0%) |
| Male | 11,357 (40.9%) | 1371 (28.7%) |
| **Niacin** | 2,185 (3.5%) | 134 (1.1%) |
| Female | 856 (2.4%) | 66 (0.9%) |
| Male | 1,329 (4.8%) | 68 (1.4%) |
| **Fibrates** | 3,058 (4.8%) | 166 (1.3%) |
| Female | 1,298 (3.7%) | 75 (1.0%) |
| Male | 1,760 (6.3%) | 91 (1.9%) |

\*Adjusted for statin, niacin, and fibrate use (see Supplementary Table 23). EA = European ancestry; AA = African ancestry; EHR = electronic health record

### European ancestry

Eleven variants met the 5 selection criteria for inclusion in our study (i.e., reported as functional, previous use as an MR proxy, previous GWAS significance, availability in BioVU, and BioVU ancestry-specific minor allele frequencies (MAF) consistent with ancestral population-wide MAF available from public databases; **Supplementary Table 2**). We then tested the association of the TG-increasing allele in these SNPs with adjusted median measured TGs to validate their use as instrumental variables. Ten variants were significantly associated with measured TGs in the EA cohort (**Supplementary Table 3**), consistent with the direction of previous reporting; there was no minor allele carriage for the remaining SNP (rs147210663) to test. We restricted subsequent analyses in the BioVU EA cohort and replication AoU cohort to those significantly associated 10 SNPs.

#### Association between validated variants and clinical phenotypes

For the primary analysis, we first examined potential associations between the 10 validated variants and 21 prespecified clinical phenotypes (**Supplementary Table 4**). We found 17 Bonferroni-corrected significant associations in BioVU patients of EA (**Figure 2; Supplementary Table 5**); there were 21 additional suggestive associations with lipid and cardiovascular phenotypes. Nine of the 10 validated variants were significantly associated with at least one of these phenotypes in the expected direction. Among the potential prespecified secondary effects, we identified suggestive associations between the TG-increasing alleles and increased risk of gout (rs1801177, OR=0.75, p=0.025), diseases of the pancreas (rs1801177, OR=1.37, p=0.024), chronic pancreatitis (rs1801177, OR=1.53, p=0.039); there was lowered risk of urinary tract infection (rs268, OR=0.88, p=0.036) and acute pancreatitis (rs268, OR=0.59, p=0.012; **Figure 2**). For the unspecified phenotypes in the global PheWAS (n=1587), there were additional associations with increased risk of cataplexy and narcolepsy (rs118204057, OR=26.84, p=1.74×10^-5^), atherosclerosis of the renal artery (rs1801177, OR=2.10, p=1.39×10^-5^), pleurisy/pleural effusion (rs1801177, OR=1.30, p=1.30×10^-5^), complication of colostomy or enterostomy (rs1801177, OR=2.20, p=1.81×10^-5^), congenital deformities of feet (rs1801177, OR=2.87, p=3.10×10^-5^), and disorders of lipoid metabolism (rs651821, OR=1.51, p=1.85×10^-5^); there was decreased risk of glaucoma (rs138326449, OR=0.35, p=1.07×10^-5^; **Supplementary Tables 6 and 7**).

**Figure 2.**
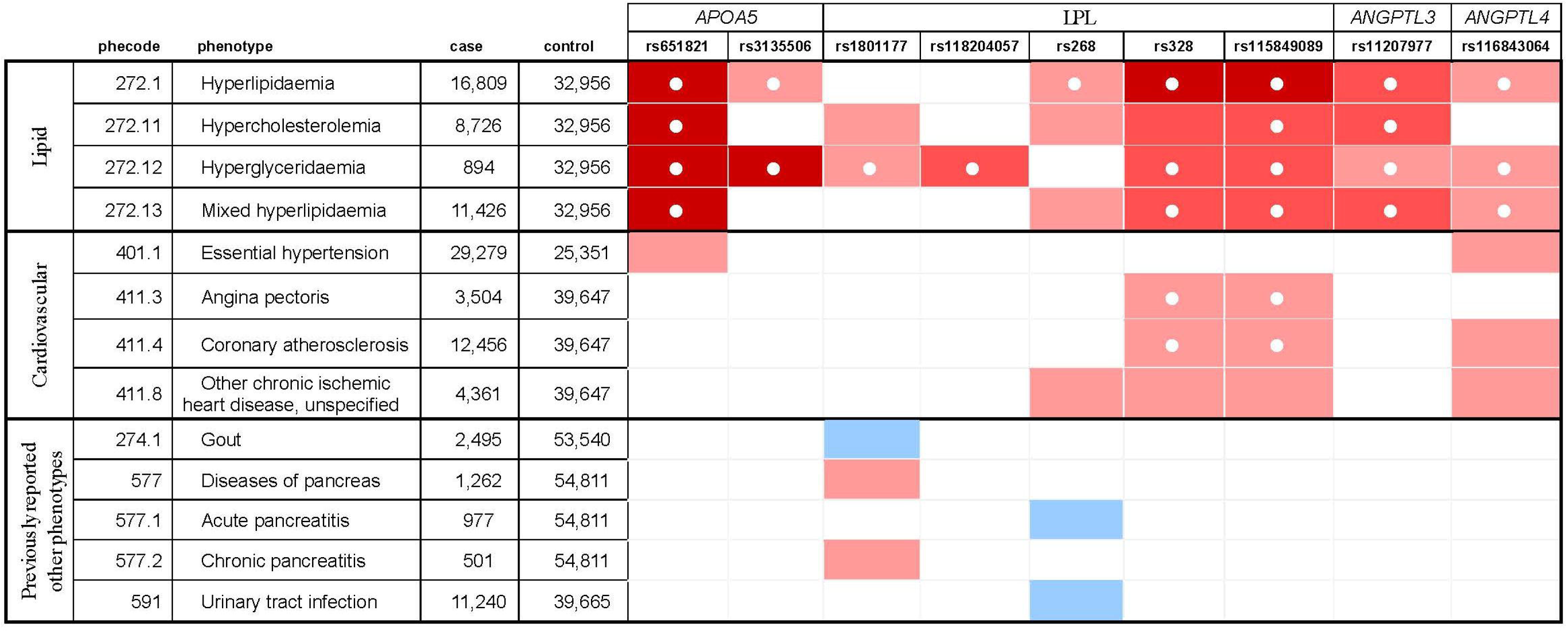
Significant associations between prespecified phenotypes and genetic variants in the BioVU European ancestry cohort, with All of Us replication indicated. ○ = replicated in All of Us. Dark red = p<3.15×10^-5^ (0.05/1587) [logistic regression]; highly significantly increased risk associated with TG-increasing allele (full PheWAS Bonferroni-corrected). Medium red = p<0.0024 (0.05/21) [logistic regression]; significantly increased risk associated with TG-increasing allele (Bonferroni-corrected). Light red = p<0.05 [logistic regression]; suggestive significance of increased risk associated with TG-increasing allele. Light blue = p<0.05 [logistic regression]; suggestive significance of decreased risk associated with TG-increasing allele.

#### Replication

In the AoU EA cohort, the validated variants (n=10) had MAF that were consistent with ancestral population wide MAF (**Supplementary Table 8**). For the 21 prespecified phenotypes, we replicated 26 significant and suggestive associations (p<0.05) with these variants (**Figure 2**; **Supplementary Table 9**). Consistent with the BioVU cohort of patients of EA, the AoU cohort had significant associations between prespecified lipid phenotypes and genetic variants (i.e., rs268, rs328, rs651821, rs3135506, rs11207977, rs115849089, rs1801177, rs118204057, and rs116843064). Additionally, we replicated associations with prespecified cardiovascular phenotypes, including angina pectoris (rs328 and rs115849089) and coronary atherosclerosis (rs328 and rs115849089). No other prespecified or unspecified phenotypes identified in the BioVU EA cohort were replicated in the AoU EA cohort (**Supplementary Tables 6 and 9**).

#### Secondary analyses

While some of the significant associations from the primary analysis were attenuated after the additional adjustment for LDL-C, results remained largely consistent (**Supplementary Table 10**). For the genetic variant set-based tests (i.e., locus-based analyses), we grouped SNPs from *LPL* and *APOA5*, with no notable additional results (**Supplementary Table 11**). For the interactions, we conducted interaction analyses for lipid and cardiovascular phenotypes among validated variants in four TG-associated genes—*APOA5* (rs651821), *LPL* (rs328), *ANGPTL3* (rs11207977), and *ANGPTL4* (rs116843064)—in the BioVU EA cohort using logistic regression models adjusted for the same covariates as the primary analysis. No statistically significant interactions were observed between any of the variant pairs. Likelihood ratio tests further confirmed that including interaction terms did not significantly improve model fit (**Supplementary Table 12**). For predicted gene expression, we found that 4 TG-associated genes had sufficient tissue samples to conduct cross-tissue analysis (*APOA5* only had a single qualifying tissue). Of these, both *LPL* and *ANGPTL3* yielded significant associations with prespecified lipid phenotypes; *LPL* also had suggestive associations with prespecified cardiovascular phenotypes (**Table 2; Supplementary Table 13**). Among the prespecified other phenotypes, *ANGPTL3* had suggestive associations with gout and migraine, while *APOC3* had a suggestive association with dementias. Single-tissue analyses did not yield any additional significant results (**Supplementary Table 14**).

**Table 2.** Significant associations between prespecified phenotypes and genetically predicted expression of TG-associated genes among BioVU patients of EA.

|  |  |  | *LPL* |  | *APOC3* |  | *ANGPTL3* |  |
| --- | --- | --- | --- | --- | --- | --- | --- | --- |
| Phenotype group | Phecode | Phenotype | GBJ_Statistic | p-value | GBJ_Statistic | p-value | GBJ_Statistic | p-value |
| Lipid phenotypes | 272.1 | Hyperlipidemia | **5.9657** | **0.001\*\*** | 1.7557 | 0.087 | **5.5513** | **0.002\*\*** |
|  | 272.11 | Hypercholesterolemia | **2.4444** | **0.045\*** | 0.7635 | 0.248 | **4.7191** | **0.004\*** |
|  | 272.12 | Hyperglyceridemia | **5.9906** | **0.001\*\*** | 0.5192 | 0.328 | **2.1245** | **0.049\*** |
|  | 272.13 | Mixed hyperlipidemia | **4.5951** | **0.005\*** | 1.5638 | 0.106 | **5.8732** | **0.001\*\*** |
| Cardiovascular phenotypes | 411.3 | Angina pectoris | **4.8790** | **0.004\*** | 0 | 1 | 0 | 1 |
|  | 411.4 | Coronary atherosclerosis | **3.4456** | **0.016\*** | 0.1981 | 0.486 | 0.0578 | 0.490 |
|  | 411.8 | Other chronic ischemic heart disease, unspecified | **4.0202** | **0.009\*** | 0 | 1 | 0 | 1 |
| Previously reported other phenotypes | 274.1 | Gout | 0.1353 | 0.557 | 0 | 1 | **4.1693** | **0.006\*** |
|  | 290.1 | Dementias | 0 | 1 | **3.4496** | **0.016\*** | 0 | 1 |
|  | 340 | Migraine | 0 | 1 | 0 | 1 | **2.4559** | **0.035\*** |

\*\*p<0.0024 (0.05/21); significant association with genetically predicted expression of gene (Bonferroni-corrected)
\*p<0.05; suggestive significance of association with genetically predicted expression of gene
bold=at least suggestive significance; red=Bonferroni-corrected significant
EA = European ancestry; GBJ = Generalized Berk Jones

### African ancestry

The same eleven variants met the 5 selection criteria for inclusion in our study (i.e., reported as functional, previous use as an MR proxy, previous GWAS significance, availability in BioVU, and BioVU ancestry-specific MAF consistent with ancestral population-wide MAF available from public databases; **Supplementary Table 2**). However, while all associations were consistent with the direction of previous reporting (**Supplementary Table 3**), only 6 of these variants were significantly associated with measured TGs; 5 variants were not associated, and there was no minor allele carriage for the remaining SNP (**Supplementary Table 3**).

#### Association between validated variants and clinical phenotypes

In the BioVU AA cohort, there were fewer significant associations among known TG-associated variants. We identified 1 suggestive association between the 6 validated TG-increasing variants and the prespecified lipid or cardiovascular clinical phenotypes: mixed hyperlipidemia (rs11207977, OR=1.14, p=4.08×10^-3^; **Figure 3**; **Supplementary Table 15**). Additionally, among prespecified other phenotypes, we found suggestive associations with lower risk for dizziness (rs11207977, OR=0.89, p=0.023) and increased risk for Alzheimer’s disease (rs651821, OR=1.57, p=0.025). For the 1065 unspecified phenotypes in the global PheWAS, there were significant associations with increased risk for heart valve replacement (rs651821, OR=1.79, p=1.36×10^-5^), hemorrhagic disorder due to intrinsic circulating anticoagulants (rs651821, OR=2.58, p=3.89×10^-6^), and decreased libido (rs1801177, OR=3.68, p=5.44×10^-6^), as well as decreased risk for stricture of artery (rs147210663, OR=0.08, p=2.00×10^-5^) and insect bite (rs147210663, OR=0.09, p=1.56×10^-5^; **Figure 3; Supplementary Tables 16 and 17**). No associations replicated in the AoU AA cohort (**Supplementary Tables 16 and 18**).

**Figure 3.**
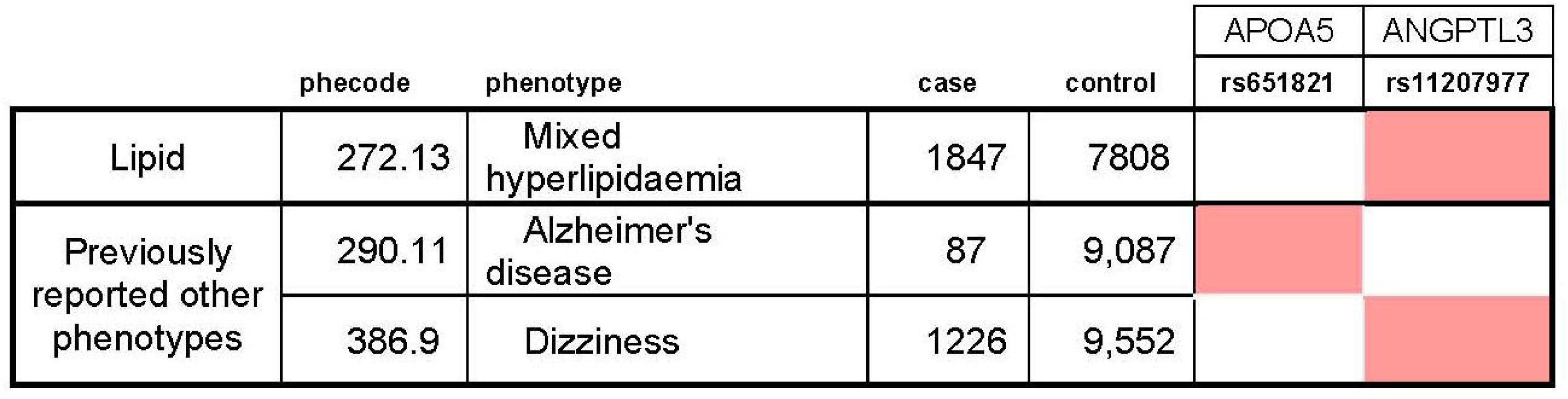
Significant associations between prespecified phenotypes and genetic variants in the BioVU African ancestry cohort, with All of Us replication indicated. ○ = replicated in All of Us. Dark red = p<4.69×10^-5^ (0.05/1065) [logistic regression]; highly significantly increased risk associated with TG-increasing allele (full PheWAS Bonferroni-corrected). Medium red = p<0.0024 (0.05/21) [logistic regression]; significantly increased risk associated with TG-increasing allele (Bonferroni-corrected). Light red = p<0.05 [logistic regression]; suggestive significance of increased risk associated with TG-increasing allele. Light blue = p<0.05 [logistic regression; suggestive significance of decreased risk associated with TG-increasing allele.

#### Secondary analyses

There were no substantive differences in the primary results after the additional adjustment for LDL-C (**Supplementary Table 19**) or locus-based analyses (**Supplementary Table 20**). There were no results consistent across genes for interaction testing. Leveraging the independent GLGC[50–55] TG GWAS AA cohorts, we identified 12 instrumental variables for TGs (**Supplementary Table 21**), 2 of which were included among the variants of the primary analysis (i.e., rs116843064 in *ANGPTL4* and rs3135506 in *APOA5*). In the BioVU AA cohort, only 7 of these variants were associated with measured TGs (**Supplementary Table 21**); when testing these 7 variants with the 21 prespecified phenotypes, we found largely consistent patterns of limited pleiotropic associations (**Supplementary Table 22**), with only a single significant result among lipid phenotypes (mixed hyperlipidemia and rs35529421, OR=1.16, p=7.99×10^-4^) and a single significant result among cardiovascular phenotypes (angina pectoris and rs10096633; OR=1.38, p=1.68×10^-4^).

## DISCUSSION

In this study, we applied PheWAS approaches to MR-identified variants, paralleling previous work to interrogate drug efficacy and adverse effects,[8,56–59] to assess the potential for both deleterious and beneficial effects of targeting TG-associated genes in populations of both EA *and* AA using two extensive EHR databases: BioVU and AoU. We found that for individuals of EA, the lipid and cardiovascular benefits were largely confirmed, while we detected few secondary effects (either previously identified or unknown) that might pose increased risk by targeting these TG-associated genes. Among individuals of AA, however, we found only limited evidence for lipid and cardiovascular benefits; as with individuals of EA, there were few secondary effects associated with these genes. The secondary analyses were largely consistent with the primary findings.

Our results support current efforts to target TG-associated genes[60,61] for the prevention and treatment of heart disease and were consistent with previous findings,[37–39] particularly among individuals of EA. Indeed, all 9 validated variants had suggestive or significant association with lipid phenotypes in the expected direction in the BioVU cohort, and all of these had at least one hyperlipidemia phenotype validated in the AoU cohort. Moreover, 6 of the validated variants had suggestive associations with heart disease-related phenotypes in at least one cohort in the expected direction; 5 variants had suggestive significance in both cohorts (although the phenotypes did not replicate in all instances). In contrast, for individuals of AA, the associations between genetic variants and hyperlipidemia-related phenotypes were more limited, consistent with previous findings.[46,62] Only 6 out of 11 previously reported variants showed significant associations with adjusted measured TG levels; of these, only one of the variants had a significant association with hyperglyceridemia (phecode 272.12) in either the BioVU or AoU cohort, and two variants showed suggestive significance; none of these results were replicated in both cohorts. None of the 6 validated SNPs were significantly associated with other cardiovascular phenotypes, although one SNP showed suggestive significance in the AoU cohort. Notably, while analysis for the IVs identified in independent GLGC AA cohorts yielded some additional significant and suggestive results in the BioVU AA cohort, these findings were limited. Given that individuals of AA tend to have lower average TG levels and lower frequencies of severe or moderate hypertriglyceridemia compared to individuals of EA,[46] it remains unclear whether the same variants have comparable effect sizes on the cardiovascular outcomes in populations of AA and EA.

Along with the consideration of efficacy for cardiovascular benefits, it was also important to consider the other potential long-term benefits and risks of these drugs; although the limited studies investigating the impact of TG-lowering therapies on non-cardiovascular phenotypes associated with TG candidate genes found no additional effects, these studies focused on European-based populations of EA.[38,39]

Pancreatitis, diabetes, and dementia/Alzheimer’s disease were among the most serious conditions previously associated with TGs and TG-related genes. Previous findings have indicated increased risk of pancreatitis with increased risk of high triglycerides;[49,63–68], our analysis of pancreatitis phenotypes among individuals of EA yielded limited results, with two SNPs (rs1801177 and rs3135506) showing suggestive significance for increased risk of related phenotypes in either the BioVU or AoU EA cohorts (not replicated), and one SNP showing suggestive significance of decreased risk (rs268). Similarly, results regarding potential benefits for diabetes prevention were not conclusive, despite previous research indicating this possibility.[35,37,69–73] Likewise, despite prior findings indicating potential linkages between TG-associated genes and ADRD, we found limited evidence of robust associations (i.e., secondary analysis of predicted gene expression found a suggestive association between dementias and *APOC3* in patients of EA) which supports ideas that TG-lowering treatment could reduce the risk of dementia or Alzheimer’s diseases.[74–78] The lack of consistent results parallel research assessing the relationship between LDL-C lowering treatments (i.e., statins) and dementias; indeed, the association between lipid management and the benefit for Alzheimer’s disease remains controversial.[79–84] We found no consistent evidence to support these potential secondary beneficial effects among individuals of AA.

Our study featured several notable strengths. First, application of PheWAS to MR-identified variants enabled an unbiased investigation of over 1,000 clinical phenotypes. Second, leveraging longitudinal EHR data from BioVU and AoU allowed us to evaluate long-term effects in individuals of EA and AA. This is particularly significant as populations of AA have historically been underrepresented in genetic and pharmacogenetic research. Moreover, the use of two large cohorts offered more robust results. Finally, by focusing on previously reported variants, our study contributed to a more nuanced understanding of the long-term effects—both beneficial and deleterious—of TG-lowering medications.

However, there were limitations. First, despite conducting analyses in two large biobanks, the cohorts of AA remained comparatively small, which limited the statistical power compared to individuals of EA, especially for low-frequency variants. Indeed, in post-hoc power calculations using PS[85,86] (with a Type 1 error=0.05 and uncorrected chi-squared statistics), given an OR=1.5 for a phenotype in the exposed subjects (as observed in the association between rs1801177 and the prespecified phenotype chronic pancreatitis [phecode 577.2] in individuals of EA), we would need 473 cases and 4730 controls (assuming a 1:10 case:control ratio) to detect an association with a genetic variant of MAF 0.05 (10% carriers), and 4067 cases and 40,670 controls to detect association with a genetic variant of MAF 0.005 (1% carriers). Likewise, given an OR=1.28 for a phenotype in the exposed subjects (as observed in the association between rs268 and the prespecified phenotype other chronic ischemic heart disease [phecode 411.8] in individuals of EA), we would need 1384 cases and 13,840 controls (assuming a 1:10 case:control ratio) to detect association with a genetic variant of MAF 0.05 (10% carriers), and 12,190 cases and 121,900 controls to detect association with a genetic variant of MAF 0.005 (1% carriers). These calculations confirm that several associations identified in EA cohorts would be underpowered for detection in the AA cohorts, particularly for variants with low allele frequency or modest effect size. These observations also apply to other ancestral populations with smaller proportions in BioVU and AoU (e.g., individuals of Asian ancestry). Future validation studies using larger, multi-ancestry cohorts are needed. As AoU continues to recruit participants, this analysis may be more informed with an updated dataset.

Second, this study focused exclusively on previously reported variants within candidate TG-lowering targets which have been identified in cohorts of EA. Given that there are differences in genetic architecture and gene-environment interactions between populations of EA and AA (e.g., effect sizes, minor allele frequences, and LD structure),[87,88] these factors, combined with cohort size, could have hampered replication of results in the AA cohorts (relative to the findings among the EA cohorts). To address this limitation, we tested instrumental variables identified in an independent AA cohort, and the results were comparable to the primary analysis (with extremely limited associations). Notably, most of the identified IVs would not pass the variant selection criteria for the primary analysis; only 7 variants were associated with measured TGs, and of these, only 1 was previously identified as a functional variant (i.e., rs3135506, a variant which overlapped with our primary analysis). Moreover, we did not perform ancestry-specific fine-mapping or trans-ancestry colocalization, which would be required to comprehensively identify causal variants in populations of AA; while we agree this analysis could be informative for identifying additional variants for targeting, this analysis fell outside the scope of our study. Further, there may be other unidentified functional variants whose impact on the association results remain unknown, particularly in populations of AA. Given the historical underrepresentation of populations of AA and other ancestral groups in genetic studies, it is possible that some important ancestry-specific TG-related variants remain unidentified. With the expansion of multi-ancestry biobanks and whole-genome sequencing data as well as the application of additional drug-targeted MR approaches, future studies could more thoroughly assess diverse ancestry groups, including investigation of low-frequency genetic variants, identification of additional targets, and identification of drug-repurposing candidates.

Third, our study cohorts were composed of patients who have survived to receive clinical care and to be included in EHR-linked biobanks. This introduces the possibility of Type 1 selection bias—particularly if the genetic variants under study influence both disease risk and survival. Variants with beneficial cardiometabolic effects may be overrepresented among older or healthier patients, potentially attenuating associations or obscuring deleterious effects. However, other than Mendelian disease and well-recognized, strongly pathogenic variants, the individual effects of most genetic variations, particularly common genetic variations, on survival is likely to be small and seem unlikely to lead to important depletion of susceptibles in a population spanning a broad age group.[7] Moreover, BioVU offers an opportunity to follow at least some patients from birth, with records spanning more than 30 years. While this follow-up may not completely mitigate issues of survivorship bias, it can help attenuate such concerns.

Fourth, additional factors (e.g., comorbidities and social determinants of health) could be unmeasured confounders; moreover, these factors could help explain why lipid-lowering effects do not correspond automatically to reduced cardiovascular risk. Notably, current efforts are underway (e.g., AoU) to systematically collect data regarding lifestyle, healthcare access and utilization, emotional health, behavioral health, and social factors of health. As more comprehensive data become available, we will have greater opportunities to integrate lifestyle, sociodemographic, and environmental covariates in future analyses.

Fifth, such factors combined with differences between the data sources (e.g., record-keeping practices and patterns of healthcare utilization) could limit our ability to replicate findings. Specifically, while both BioVU and AoU include large, EHR-linked populations, some differences (e.g., age distribution, geographic region, healthcare delivery systems, and duration of EHR follow-up) could influence phenotype prevalence, comorbidity patterns, or data completeness, which could in turn affect statistical power and reproducibility.

Sixth, we were unable to complete analysis of *APOA5* in the predicted expression secondary analysis, given the limited availability of tissues. Moreover, the limited availability of ancestry-diverse expression quantitative trait loci (eQTL) resources impeded our ability to conduct predicted expression analyses for the BioVU and AoU AA cohorts. The ongoing development of such resources[89–92] can facilitate future ancestry-matched transcriptome-wide association studies.

Seventh, detecting protective effects can be more challenging than demonstrating adverse effects related to known risk alleles (e.g., lowered diabetes risk). It is possible that there will be protective benefits from drugs targeting TG-lowering genes, but we do not yet have sufficient power to detect these effects.

Finally, while the combined MR-PheWAS approach was designed to generate hypotheses regarding vertical pleiotropic effects of functional variants, we note that alterations in the selected TG-related genes may lead to additional clinical consequences via causal pathways that are not solely affected by TGs—namely, horizontal pleiotropic effects—because the candidate genes under examination may have biological functions beyond TG regulation. In particular, as noted in the Introduction, TGs are closely related to other atherogenic lipids, including LDL-C and ApoB, which are well-established causal factors for CHD. Horizontal pleiotropy can complicate clinical interpretation (e.g., biasing effect estimates) and parsing the relationships remains important; indeed, recent multivariable MR studies suggest that associations between TG and CHD were attenuated after adjusting for ApoB-containing lipoproteins, rather than TG per se. Nevertheless, our findings remain relevant for evaluating the clinical consequences of modulating these genes by focusing on the detection of potential effects of the TG-related pathways, especially as several therapeutics targeting these genes have been approved (e.g., evinacumab, an ANGPTL3 inhibitor) or are currently in development. Most importantly, this analysis was not intended to be the conclusive determination of such effects, but rather an important hypothesis-generating component of the larger project of assessing such potential effects; future work should include testing potential alternative pathways as well as monitoring effects in populations using TG-targeting therapies.

In conclusion, by leveraging two large EHR-based biobanks (BioVU and AoU), we applied genetic approaches to evaluate the associations between clinical phenotypes and previously reported TG variants in populations of EA and AA. We confirmed the associations with TG-related phenotypes for most of the candidate genes in individuals of EA; these effects could have implications for preventing cardiovascular disease. Our results also suggest the potential for a few additional beneficial effects, with few deleterious effects, for individuals of EA using TG-lowering treatments; however, these results do not provide comparable evidence to assess the efficacy and safety of these treatments for individuals of AA. As datasets continue to grow in breadth (e.g., whole genome sequencing and survey data regarding social drivers of health) and length of follow-up, testing in populations other than those of EA is a priority.

## METHODS

The primary analysis was conducted in ancestry-specific cohorts of patients of EA and AA from BioVU, a biobank at VUMC. BioVU offers genome-wide genotyping data for ∼90,000 patients on the Illumina Infinium Expanded Multi-Ethnic Genotyping Array plus custom content platform (MEGA), which are linked to de-identified copies of the patients’ EHRs. These EHRs include demographics, clinical notes, lab results, medications, and diagnostic and procedure codes (e.g., International Classification of Diseases, Ninth Revision, Clinical Modification [ICD-9-CM] codes and Tenth Revision [ICD-10-CM] codes).[93] Replication analysis was conducted among comparable cohorts of participants of EA and AA in AoU (**Supplementary §1**). This study was considered non-human subjects and was approved by the VUMC Institutional Review Board (IRB# 220725).

### Cohort Eligibility

Primary cohort eligibility criteria included BioVU patients with available MEGA genotyping data that passed standard quality controls (see below; **Supplementary §2**), and at least 2 ICD codes recorded in their EHR through study end date (January 11, 2023). We restricted the study to patients with genetically-determined predominantly EA or AA).[94] Although socially identified EHR-reported race is largely consistent with genetic ancestry in BioVU,[95–97] to minimize the conflation of genetic and social factors, we further restricted the cohorts to patients with consistent EHR-reported race (i.e., White race for patients of EA and Black race for patients of AA); individuals with more than one reported race were excluded. We further included adjustment for the first 10 principal components (PCs) to help account for intra-ancestral variability.

### Clinical characteristics

Patient characteristics were extracted from the EHR, including age (at most recent EHR visit), EHR-reported gender, length of EHR (i.e., the total time between an individual’s first and most recent EHR record through the end of the study), lipid levels (i.e., TG, HDL-C, and LDL-C), and whether patients were ever prescribed lipid-lowering medications (statins, niacin, or fibrates; **Supplementary Table 23**); median lipid values were calculated using all measurements after quality control (excluding those that were biologically implausible, e.g. negative values). We adjusted median TG levels for use of lipid-lowering medications for each individual.[46,98]

### Genotyping and Imputation

Genotyping was performed on the MEGA platform with quality control as performed previously (**Supplementary §2**).[99–101] Genetically-determined ancestry and the first 10 PCs for ancestry were determined following previous reports.[94]

### Genetic variants in TG gene targets

#### Primary analysis

We assessed variants selected from 5 genes that have been identified previously by MR as TG-lowering targets: *APOA5*, *LPL*, *APOC3*, *ANGPTL3*, and *ANGPTL4* (**Supplementary Table 2**).[37,49] To be included in the analysis, variants were required to demonstrate the following: (1) relevant function (i.e., protein-coding missense variants or variants known to influence gene expression); (2) prior use as proxies in MR studies to test associations between target genes and clinical phenotypes other than TG levels; (3) availability in BioVU; (4) ancestry-specific previously-reported significant associations (p < 5×10^−^□) in large-scale GWAS; and (5) consistent ancestry-specific MAF with 1000 Genomes reference data (**Figure 1**).[102] For SNPs meeting these criteria, we then assessed whether the TG-increasing allele of the genotype was significantly associated with adjusted measured TG levels in the direction previously reported in separate analyses of the EA and AA BioVU cohorts; further analyses testing associations with phenotypes (see below) were restricted to these TG-increasing variants significantly associated with measured TG levels (p<0.05) in the expected direction.

#### Secondary analyses

We conducted the following secondary analyses:

*1) Additional measured LDL-C adjustment*. While elevated TG levels are consistently associated with increased CHD risk, disentangling their independent causal role remains complex due to strong correlations with other lipid traits, particularly ApoB and LDL-C. As such, we conducted analyses in the BioVU EA and AA cohorts with an additional adjustment for median LDL-C levels. As ApoB levels are not routinely collected in clinical practice, we could not additionally adjust for ApoB.
*2) Genetic variant set-based tests*. We used the Generalized Berk-Jones (GBJ) test to analyze the validated variants in the same locus together for increased power in the BioVU EA and AA cohorts.
*3) Predicted gene expression*. We conducted a secondary analysis testing the association between the prespecified phenotypes (see below) and the predicted gene expression of the 5 TG-lowering genes; we restricted this analysis to individuals of EA, since most eQTL-based gene expression prediction models—such as those we used from GTEx v8—are largely derived from European ancestry populations[103] and may be less accurate for individuals of other ancestral groups.[104–106] Selecting tissues for availability and biological relevance, we assessed cross-tissue effects relative to the 21 prespecified phenotypes, using (1) kidney and liver tissues for *ANGPTL3*; (2) tibial atrial, kidney, and heart atrial tissues for *ANGPTL4*; (3) pancreatic tissue for *APOA5*; (4) heart left ventricular and kidney tissues for *APOC3*; and (5) kidney and whole blood tissues for *LPL*. We prioritized cross-tissue analyses because most drug effects are systemic and may not be limited to a single biologically “relevant” tissue; further, this approach accounts for potential correlation between tissues and enhances power to detect consistent effects while reducing false positives from single-tissue noise. However, we also conducted single tissue analyses to improve interpretability.
*4) Interaction in significant results*. We conducted analyses assessing interactions between variants for phenotypes with significant associations in more than one gene within a BioVU cohort.
*5) Instrumental variables identified by querying an independent AA cohort*. Because the functional variants associated with TGs have been identified among cohorts of European ancestry, we conducted an additional analysis to examine instrumental variables in the 5 TG-related genes derived from a world-wide, multi-institution cohort of individuals with AA. We leveraged the GLGC[50–55] TG GWAS within AA cohorts and selected AA-specific IVs for TGs. Specifically, we selected variants within +/- 100kb of the candidate gene regions which were associated with measured TG levels at genome-wide significance (p<5×10^-8^) in the GLGC AA cohorts. We also conducted clumping to select independent variants (R^2^<0.01). Among the identified IVs, we assessed whether the variants were associated with measured TGs in the BioVU AA cohort; for the validated variants, we assessed associations with the 21 preselected phenotypes (see below) in the BioVU AA cohort.

### Phenotypes

#### Phecodes

We established the presence or absence of clinical phenotypes using phecodes, a system based on clinical diagnosis codes (i.e., ICD-9-CM and ICD-10-CM).[56,57] Cases were individuals with 2 or more occurrences of the phecode; controls had zero occurrences; and individuals with 1 mention of the code or related codes were excluded from the analysis to limit misclassification. We required ≥50 cases to include a phecode for analysis.

#### Phenotype categories

We separated the qualifying phecodes (i.e., ≥50 cases) into 4 categories for analysis in both cohorts, based on previous evidence of association with measured TGs or genetically-determined TG levels (**Supplementary Table 4**): (1) 4 prespecified lipid phenotypes [i.e., hyperlipidemia, hypercholesterolemia, hyperglyceridemia, and mixed hyperlipidemia], to confirm the efficacy of the target; (2) 4 prespecified cardiovascular phenotypes [i.e., essential hypertension, angina pectoris, coronary atherosclerosis, and other chronic ischemic heart disease, unspecified], to confirm the desired secondary cardiovascular benefits; (3) 13 prespecified other phenotypes previously associated with TGs [i.e., type 2 diabetes, gout, dementias, Alzheimer’s disease, vascular dementia, delirium due to conditions classified elsewhere, headache, migraine, dizziness, diseases of pancreas, acute pancreatitis, chronic pancreatitis, and urinary tract infection], to determine if reduced lipid or cardiovascular risk could coincide with elevated risk for known potential secondary outcomes; and (4) all remaining qualifying phecodes in a global PheWAS, to assess whether there are unknown outcomes associated with targeting TG-lowering genes.

### Statistical Analysis

We used linear regressions adjusted for age and sex to assess whether qualifying genotypes were significantly associated with adjusted measured TG levels in separate analyses of the EA and AA BioVU cohorts.

For the primary analysis among the EA and AA cohorts in BioVU and AoU, we used logistic regression to test the associations between phecodes and validated TG-increasing variants, with adjustment for age at most recent clinical visit, sex, length of EHR, and the first 10 PCs for ancestry. In BioVU, for associations with the prespecified clinical phenotypes (i.e., categories 1-3; n=21), we considered p<0.05 as suggestive, p<0.0024 (0.05/21) as significant (Bonferroni-corrected to account for multiple testing), and p<3.15×10^-5^ and p<4.69×10-5 (Bonferroni-corrected significance for the global PheWAS, as below) as highly significant in the EA and AA cohorts, respectively; we also report significance (p<0.05) corrected for false discovery rate (FDR) for greater interpretability of the results. For the remaining phenotypes in BioVU we considered p<3.15×10^-5^ (0.05/1587 unspecified phecodes tested) and p<4.69×10^-5^ (0.05/1065 unspecified phecodes tested) as significant in patients of EA and AA, respectively. For the AoU replication cohorts, p<0.05 was considered significant.

In the secondary analysis, 1) for the additional LDL-C adjustment, we used logistic regression adjusted for median LDL-C, age at most recent clinical visit, sex, length of EHR, and the first 10 PCs for ancestry. We considered p<0.0024 (0.05/21) as significant and p<0.05 as suggestive for these tests. 2) For genetic variant set-based tests, we used the GBJ test[107] to analyze the variants in the same locus for increased power in the BioVU EA and AA cohorts. We considered p<0.0024 (0.05/21) as significant and p<0.05 as suggestive for these tests. 3) For interactions, in selected phenotypes, we conducted a logistic regression for selected variants from genes with significant results, adjusted for age at most recent clinical visit, sex, length of EHR, and the first 10 PCs for ancestry (Model 2). We additionally adjusted for variant-pair interactions (Model 1). We then conducted likelihood ratio tests between these models; we considered p<0.05 as significant for these tests. 4) For the predicted gene expression, comparing PrediXcan, UTMOST, and JTI methods,[34,108,109] we identified the best performing gene expression model for each gene by selecting the model with the highest performance R^2^ (see SNPs for each tissue in **Supplementary Table 24**). Then, we assessed the cross-tissue effects between (1) the predicted gene expressions for each gene with more than one available tissue and (2) the 21 pre-specified clinical phenotypes (**Supplementary Table 4**), using the GBJ test[107] to account for potential expression correlation among the different tissues.[110,111] We considered p<0.0024 (0.05/21) as significant and p<0.05 as suggestive for these tests. We also conducted tissue-specific analyses to provide context for interpreting the cross-tissue results; for single-tissue results, we used logistic regressions to test the associations between the 21 prespecified phenotypes and the predicted expression for each tissue, adjusted for age at most recent clinical visit, sex, length of EHR, and the first 10 PCs for ancestry. And 5) for the instrumental variables identified in an independent GLGC AA cohort, we used we used logistic regression adjusted for age at most recent clinical visit, sex, length of EHR, and the first 10 PCs for ancestry.

Individuals with missingness for any covariate were omitted from primary and secondary analyses. We present categorical variables as numbers and percentages and continuous variables as means and standard deviation (SD). All analyses were conducted using R version 4.1.0 and PLINK version 2.0.

## Data Availability

All data produced in the present study are available upon reasonable request to the authors.

https://github.com/FengLabVUMC/Triglyceride-PheWAS

## List of abbreviations

AA: African ancestry
ADRD: Alzheimer’s disease and related dementias
AoU: All of Us Research Program
ApoB: apolipoprotein B
CHD: coronary heart disease
EA: European ancestry
EHR: electronic health record
eQTL: expression quantitative trait loci
FDR: false discovery rate
GBJ: generalized Berk-Jones
GLGC: Global Lipids Genetics Consortium
HDL-C: high-density lipoprotein cholesterol
ICD-9-CM: International Classification of Diseases, Ninth Revision, Clinical Modification
ICD-10-CM: International Classification of Diseases, Tenth Revision, Clinical Modification
IRB: Institutional Review Board
LDL-C: low density lipoprotein cholesterol
MAF: minor allele frequencies
MEGA: Illumina Infinium Expanded Multi-Ethnic Genotyping Array
MR: Mendelian randomization
PC: principal component
PheWAS: phenome wide association study
SD: standard deviation
TG: triglyceride
VUMC: Vanderbilt University Medical Center

## DECLARATIONS

### Ethics approval and consent to participate

The study was approved by the VUMC Institutional Review Board (IRB# 220725).

### Availability of data and materials

#### BioVU data and code

Statistical code: Available at https://github.com/FengLabVUMC/Triglyceride-PheWAS. Data set: Full summary statistics for validated variants and qualifying phenotypes for the BioVU cohorts are available in file Supplementary Tables 7-24. For individual level data, VUMC EHR data are de-identified using Safe-harbor methods. BioVU genomic data are linked to de-identified records and are further protected by BioVU data use agreements ensuring that researchers will not attempt re-identification. BioVU participant consenting procedures limit access to individual level data. Limited primary cohort data are available by request to Dr. Feng (e-mail), pending BioVU approval and a data use agreement.

#### All of Us data and code

In line with the privacy standards set by the All of Us Research Program, data and code used for these cohorts are available to approved researchers who register for access to the Researcher Workbench platform at https://workbench.researchallofus.org/login. This analysis was run on the All of Us Research Program Controlled Tier Dataset version 7, production release C2022Q4R9.

### Funding acknowledgements

R01HL163854 (Q.F.), R01GM120523 (Q.F.), R01HL133786 (W.Q.W.), R01HL171809 (Q.F, W.Q.W) and Vanderbilt Faculty Research Scholar Fund (Q.F.).

The primary dataset(s) were obtained from Vanderbilt University Medical Center’s BioVU, which is supported by institutional funding, 1S10RR025141-01, and CTSA grants UL1TR002243, UL1TR000445, and UL1RR024975. Additional funding provided by the NIH through grants P50GM115305 and U19HL065962. The authors wish to acknowledge the expert technical support of the VANTAGE and VANGARD core facilities, supported in part by the Vanderbilt-Ingram Cancer Center (P30 CA068485) and Vanderbilt Vision Center (P30 EY08126).

The validation dataset(s) were obtained from the All of Us Research Program. This program would not be possible without the partnership of its participants. Additionally, the All of Us Research Program is supported by the National Institutes of Health, Office of the Director: Regional Medical Centers: 1 OT2 OD026549; 1 OT2 OD026554; 1 OT2 OD026557; 1 OT2 OD026556; 1 OT2 OD026550; 1 OT2 OD 026552; 1 OT2 OD026553; 1 OT2 OD026548; 1 OT2 OD026551; 1 OT2 OD026555; IAA #: AOD 16037; Federally Qualified Health Centers: HHSN 263201600085U; Data and Research Center: 5 U2C OD023196; Biobank: 1 U24 OD023121; The Participant Center: U24 OD023176; Participant Technology Systems Center: 1 U24 OD023163; Communications and Engagement: 3 OT2 OD023205; 3 OT2 OD023206; and Community Partners: 1 OT2 OD025277; 3 OT2 OD025315; 1 OT2 OD025337; 1 OT2 OD025276.

### Contributors

Conceptualization (ALD, WQW, CMS, QF); data curation (EO, YX, ALD, SM, RT, GK, SS, LJ, NJC, WQW, QF); formal analysis (EO, YX); investigation (EO, YX, ALD, SM, RT, XZ, CMS, QF); funding acquisition (WQW, QF); methodology (EO, YX); validation (YX, ALD, SM, WQW, CMS, QF); project administration (ALD, WQW, CMS, QF), visualization (EO, YX); supervision (ALD, WQW, CMS, QF); resources (SS, LJ, NJC, WQW, QF); writing – original draft (EO, YX, ALD, CMS, QF); and writing - review and editing (EO, YX, ALD, SM, RT, XZ, GK, SS, LJ, NJC, CMS, QF)

The first and corresponding authors had full access to all the data in the study and take responsibility for the integrity of the data and the accuracy of the data analysis. All authors read and approved the final version of the manuscript.

