## supplementary tables and files for "Assessing long-term pleiotropic effects of potential novel triglyceride-lowering medications using variants identified by Mendelian randomization"

**Table of Contents**

Supplementary Table 1: Demographic summary of All of Us cohorts

Supplementary Table 2: Genetic variants in TG-lowering targets which met 5 selection criteria

Supplementary Table 3: Associations between previously reported TG-increasing variants and measured TGs in BioVU patients

Supplementary Table 4: Clinical phenotypes previously associated with TG-lowering genes or measured TGs

Supplementary §1. All of Us

Supplementary §2. Genotyping Quality Control in BioVU

Supplementary Table 23: Lipid-lowering medications

Supplementary Table 24: SNPs used to build predicted gene expression for TG-associated genes, by tissue

**Supplementary Table 1: Demographic summary of All of Us cohorts**

|  | **EA cohort** | **AA cohort** |
| --- | --- | --- |
| **Total N** | 97,545 | 31,710 |
| **Female, n (%)** | 59,350 (60.84%) | 19,166 (60.44%) |
| **Age (mean ± SD, years)** | 59.22±16.77 | 53.16±14.58 |
| Female | 57.40±16.78 | 52.44±15.04 |
| Male | 62.05±16.35 | 54.26±13.78 |
| **Length of EHR (mean ± SD, years)** | 9.71±8.07 | 8.03±5.84 |
| Female | 9.79±8.22 | 8.70±6.15 |
| Male | 9.57±7.85 | 7.00±5.16 |
| **Lipid measures (mean ± SD, mg/dL)** | | |
| **Triglycerides*** | 138.39 ± 126.39  (n=57,298) | 123.73 ± 122.39  (n=15,041) |
| Female | 130.55 ± 107.58  (n=34,027) | 117.49 ± 114.71  (n=9,966) |
| Male | 147.43 ± 144.58  (n=23,271) | 136.17 ± 135.57  (n=5.075) |
| **LDL-C** | 93.33 ± 43.92  (n=55,264) | 88.59 ± 48.78  (n=14,323) |
| Female | 97.6 ± 45.05  (n=32,579) | 90.25 ± 49.77  (n=9,552) |
| Male | 88.5 ± 42.09  (n=22,685) | 85.13 ± 46.44  (n=4,771) |
| **HDL-C** | 71.28 ± 39.42  (n=56,118) | 60.82 ± 30.88  (n=14,876) |
| Female | 77.6 ± 39.52  (n=33,703) | 63.32 ± 30.74  (n=9,919) |
| Male | 63.69 ± 37.92  (n=22,415) | 55.71 ± 30.54  (n=4,957) |
| **Lipid-lowering medications, n (%)** | | |
| **Statins** | 33,316 (34.2%) | 9,434 (29.8%) |
| Female | 16,630 (28.0%) | 5,705 (29.8%) |
| Male | 16,686 (43.7%) | 3,729 (29.7%) |
| **Niacin** | 3,718 (3.8%) | 1,212 (3.8%) |
| Female | 1,747 (2.9%) | 849 (4.4%) |
| Male | 1,971 (5.1%) | 363 (2.9%) |
| **Fibrates** | 7,647 (7.8%) | 1,844 (5.8%) |
| Female | 3,072 (5.2%) | 968 (5.1%) |
| Male | 4,575 (12.0%) | 876 (7.0%) |

*Adjusted for statin, niacin, and fibrate use (see Supplementary Table 23).

EA=European ancestry; AA=African ancestry; EHR=electronic health record

| **Supplementary Table 2. Genetic variants in TG-lowering targets which met 5 selection criteria.** | | | | | | | | | | | | | |
| --- | --- | --- | --- | --- | --- | --- | --- | --- | --- | --- | --- | --- | --- |
| **TG-lowering targets** | **rsNumber** | **Genomic region** | **Variant type** | **Genotyped or imputation r^2^*** | **Functional annotation** | **Previous use as proxy for MR** | **BioVU availability** | **Previous GWAS with significant associations in EA cohorts** | **Previous GWAS with significant associations in AA cohorts** | **MAF in BioVU patients of EA** | **MAF in 1000G patients of EA**** | **MAF in BioVU patients of AA** | **MAF in 1000G patients of AA***** |
| ***APOA5*** | rs651821 | 11q23.3 | 5' UTR | genotyped | Regulatory variant | ^1^ | yes | ^2–4^ | ^2–4^ | 0.061 | 0.083 | 0.152 | 0.160 |
|  | rs3135506 | 11q23.3 | Coding | genotyped | Missense (p.Ser19Trp) | ^1,5^ | yes | ^6^ | ^2–4^ | 0.065 | 0.068 | 0.060 | 0.067 |
| ***LPL*** | rs1801177 | 8p21.3 | Coding | genotyped | Missense (p.Asp36Asn) | ^1,7,8^ | yes | ^9,10^ | ^2–4^ | 0.017 | 0.011 | 0.045 | 0.051 |
|  | rs118204057 | 8p21.3 | Coding | genotyped | Missense (p.Gly215Glu) | ^1,8,11^ | yes | ^12,13^ | ^9,10^ | 0.0003 | 0 | 0 | 0 |
|  | rs268 | 8p21.3 | Coding | genotyped | Missense (p.Asn318Ser) | ^1,8^ | yes | ^9,14–17^ | ^2–4^ | 0.019 | 0.014 | 0.004 | 0.001 |
|  | rs328 | 8p21.3 | Coding | genotyped | Nonsense (p.Ser474Ter) | ^1,7,8^ | yes | ^2–4^ | ^2–4^ | 0.104 | 0.130 | 0.067 | 0.061 |
|  | rs115849089 | 8p21.3 | 3' UTR | genotyped | Regulatory variant | ^18,19^ | yes | ^2–4^ | ^2–4^ | 0.117 | 0.145 | 0.045 | 0.058 |
| ***APOC3*** | rs138326449 | 11q23.3 | Splice site | 0.73 | Splice donor variant (IVS2+1G>A) | ^1,20,21^ | yes | ^12,13^ | ^2–4^ | 0.003 | 0.003 | 0.0006 | 0 |
|  | rs147210663 | 11q23.3 | Coding | genotyped | Missense (p.Ala43Thr) | ^1,20,21^ | yes | ^9,10,12,13^ | ^9,10,22^ | 0 | 0 | 0.002 | 0.009 |
| ***ANGPTL3*** | rs11207977 | 1p31.3 | Intronic | 0.99 | eQTL/Regulatory | ^18,19^ | yes | ^2–4^ | ^2–4^ | 0.338 | 0.301 | 0.330 | 0.323 |
| ***ANGPTL4*** | rs116843064 | 19p13.2 | Coding | genotyped | Missense (p.Glu40Lys) | ^1,7,23^ | yes | ^24–26^ | ^2–4^ | 0.015 | 0.026 | 0.003 | 0.003 |

TG=triglyceride, MR=Mendelian randomization, EA=European ancestry, AA=African ancestry, GWAS=genome-wide association study, MAF=minor allele frequency

* For imputed variants, we report the imputation r2 here.

** 1000 Genome EUR population

*** 1000 Genome AFR population

**Supplementary Table 3: Associations between previously reported TG-increasing variants and measured TGs in BioVU patients**

|  |  |  |  |  | **European ancestry patients**  **N=29,327** | | **African ancestry patients**  **N=5,587** | |
| --- | --- | --- | --- | --- | --- | --- | --- | --- |
| **Gene** | **SNP** | **Minor allele** | **Allele associated with increased TGs, as previously reported** | **References** | **beta** | **P** | **beta** | **P** |
| ***APOA5*** | **rs651821** | C | C | ^1^ | 0.1309 | 6.156x10^-47^ | 0.02674 | 0.0395 |
|  | **rs3135506** | C | C | ^1^ | 0.13 | 3.740x10^-47^ | 0.08133 | 4.238x10^-05^ |
| ***LPL*** | **rs1801177** | A | A | ^1^ | 0.08715 | 5.586x10^-07^ | 0.04598 | 0.0426 |
|  | **rs118204057** | A | A | ^1^ | 0.4206 | 6.649x10^-04^ | * | * |
|  | **rs268** | G | G | ^1^ | 0.1253 | 1.366x10^-15^ | 0.02549 | 0.7554 |
|  | **rs328** | G | C | ^1,27^ | 0.1066 | 3.488x10^-49^ | 0.04569 | 0.0166 |
|  | **rs115849089** | A | G | ^18^ | 0.09668 | 1.171x10^-44^ | 0.01617 | 0.4953 |
| ***APOC3*** | **rs138326449** | A | G | ^1^ | 0.3361 | 1.702x10^-17^ | 0.2729 | 0.1839 |
|  | **rs147210663** | A | G | ^9^ | * | * | 0.3205 | 2.267x10^-03^ |
| ***ANGPTL3*** | **rs11207977** | T | C | ^18^ | 0.03126 | 2.964x10^-11^ | 0.03624 | 3.075x10^-04^ |
| ***ANGPTL4*** | **rs116843064** | A | G | ^1,18^ | 0.1322 | 5.644x10^-13^ | 0.101 | 0.2430 |

TGs=triglycerides

*no minor allele carriage

red = significant association

**Supplementary Table 4: Clinical phenotypes previously associated with TG-lowering genes or measured TGs**

| **Phenotype group** | **Phecode** | **Clinical phenotypes** | **Source** |
| --- | --- | --- | --- |
| Lipid phenotypes | 272.1 | Hyperlipidemia | ^28^ |
|  | 272.11 | Hypercholesterolemia | ^28^ |
|  | 272.12 | Hyperglyceridemia | ^28^ |
|  | 272.13 | Mixed hyperlipidemia | ^28^ |
| Cardiovascular phenotypes | 401.1 | Essential hypertension | ^28^ |
|  | 411.3 | Angina pectoris | ^28^ |
|  | 411.4 | Coronary atherosclerosis | ^28^ |
|  | 411.8 | Other chronic ischemic heart disease, unspecified | ^28^ |
| Previously reported other phenotypes | 250.2 | Type 2 diabetes | ^29–32^ |
|  | 274.1 | Gout | ^28^ |
|  | 290.1 | Dementias | ^28^ |
|  | 290.11 | Alzheimer's disease | ^28^ |
|  | 290.16 | Vascular dementia | ^28^ |
|  | 290.2 | Delirium due to conditions classified elsewhere | ^28^ |
|  | 339 | Headache | ^31,33,34^ |
|  | 340 | Migraine | ^33,35–38^ |
|  | 386.9 | Dizziness | ^31,39^ |
|  | 577 | Diseases of pancreas | ^40,41^ |
|  | 577.1 | Acute pancreatitis | ^29,41–44^ |
|  | 577.2 | Chronic pancreatitis | ^41,45–47^ |
|  | 591 | Urinary tract infection | ^34,39,48^ |

**Supplementary §1. All of Us**

We validated results in cohorts of predominantly European ancestry (EA) and African ancestry (AA) individuals in the All of Us (AoU) Curated Data Depository (Controlled Tier version 7, released April 20, 2023^49^). AoU is an NIH project that aims to build the most diverse cohort in history; approximately 45% of participants are racial and ethnic minorities, and >80% of participants have been historically underrepresented in biomedical research.^50^ AoU data include EHRs, genetic information, survey results, and mobile device data as of July 1, 2022 (the cutoff for the version 7 release). All data acquisition, phenotyping, and analyses were completed in the AoU Researcher Workbench, a cloud-based working environment. We applied comparable restrictions to those of the primary cohorts to determine cohort eligibility—namely, available genetic data, ≥2 ICD codes in the EHR, and genetically-determined EA or AA ancestry. As in BioVU, although socially-identified race and genetic ancestry are largely consistent in AoU,^51^ we further restricted the cohorts to participants with White or Black socially identified race, respectively, to minimize misattributions of social and genetic factors. All genetic data, including genetically determined ancestry and variants, were extracted from AoU whole genome sequencing (WGS) data version 7. In accordance with AoU policy, we report categories with fewer than 20 individuals as N≤20, and the results of corresponding calculations (e.g., percentages) are omitted to prevent reverse calculation.

We extracted participant characteristics from the EHR, including age (at most recent EHR visit), sex, and length of EHR. We also utilized the first 10 principal components as calculated independently by AoU for adjustment in analyses.^51^

We assessed the genetic variants, as validated in the BioVU EA and AA cohorts, for the AoU EA and AA cohorts, respectively. The AoU WGS includes genetic variants with more than two alleles (e.g., rs1801177); as such, we excluded the alternative alleles with the lowest MAF (e.g., C allele for rs1801177), verified the remaining alleles matched those in BioVU, and then completed the remaining analysis with the single minor allele. We confirmed the MAFs were consistent with those reported by the 1000 Genomes project and then conducted PheWAS in the EA and AA cohorts, applying the same PheWAS criteria as for the primary cohorts of EA and AA BioVU patients, respectively.

**Supplementary §2. Genotyping Quality Control in BioVU**

Genotyping at Vanderbilt University Medical Center (VUMC) for this study was completed as part of a larger VUMC initiative; we followed quality control (QC) steps as reported previously.^52–54^ Using the Illumina Infinium® Expanded Multi-Ethnic Genotyping Array plus custom content platform (VUMC BioVU MEGA^EX^) platform to deliver dense genotyping data, VUMC BioVU paired these data with de-identified clinical records. In 2019, over 100,000 samples, including HapMap controls, were clustered and then filtered for the following reasons: lack of concordance in a HapMap Mendel/Concordance Evaluation, variant call rate<0.95, unexplained relatedness (including unexpected duplication), and discrepancies between genetic sex and reported gender. We performed the same QC steps for our cohorts.

We then completed whole genome imputation using the Michigan Imputation Server.^55^ We filtered variants with (1) low imputation quality (r^2^ <0.3), and (2) a minor allele frequency (MAF) with an absolute difference >0.3 compared to the HRC reference panel. Genetically-determined ancestry and principal components (PCs) for ancestry were determined independently from AoU using the Fast and Robust Ancestry Prediction by using Online singular value decomposition and Shrinkage Adjustment (FRAPOSA) pipeline.^56^ Applying an online data augmentation, decomposition and Procrustes (OADP) approach, which utilizes a computationally efficient online singular value decomposition algorithm, we used BioVU genetic data as study dataset and used 1000 Genomes data as the reference dataset to generate harmonized datasets. Subsequently, we computed PC embeddings by projecting the study dataset samples onto a specified number of reference PCs (dimensions=20) derived from the reference dataset. Finally, we classified study individuals into ancestral populations based on their PC scores, and output predicted ancestries along with associated probabilities and distances.

**Supplementary Table 23: Lipid-lowering medications**

| **Drug Class (TG adjustment**^57^**)*** | **Generic** | **Brand** |
| --- | --- | --- |
| Statins (+18.4 mg/dL) | simvastatin | Zocor |
|  |  | FloLipid |
|  | ezetimibe-simvastatin | Vytorin |
|  | sitagliptin-simvastatin | Juvisync |
|  | simvastatin-niacin | Simcor |
|  | fenofibrate-simvastatin | Cholib |
|  | rosuvastatin | Crestor |
|  |  | Ezallor |
|  | ezetimibe-rosuvastatin | Roszet |
|  | atorvastatin | Lipitor |
|  |  | Atorvaliq |
|  | amlodipine-atorvastatin | Caduet |
|  | ezetimibe-atorvastatin | Liptruzet |
|  | pravastatin | Pravachol |
|  | pravastatin-aspirin | Pravigard |
|  | fenofibrate-pravastatin | Pravafenix |
|  | lovastatin | Mevacor |
|  |  | Altocor |
|  |  | Altoprev |
|  | lovastatin-niacin | Advicor |
|  | fluvastatin | Lescol |
|  | cerivastatin | Baycol |
|  |  | Lipobay |
|  | pitavastatin | Livalo |
|  |  | Zypitamag |
| Niacin (+89.4 mg/dL) | niacin | Niacor |
|  |  | Niacin-50 |
|  |  | Niacin SR |
|  |  | Slo-Niacin |
|  |  | Endur-acin |
|  | nicotinic acid |  |
|  | vitamin B3 |  |
|  | nicotinamide | Niaspan |
|  | niacinamide | Endur-Amide |
|  | nicotinamide riboside | Tru Niagen |
|  | nicotinamide adenine dinucleotide |  |
| Fibrates (+57.1 mg/dL) | clofibrate | Atromid-s |
|  | bezafibrate | Bezalip |
|  | fenofibrate | Antara |
|  |  | Atorva TG |
|  |  | Fenoglide |
|  |  | Fenogal |
|  |  | Fenocor |
|  |  | Fibricor |
|  |  | Golip |
|  |  | Lipanthyl |
|  |  | Lipantil |
|  |  | Lipidil |
|  |  | Lipofen |
|  |  | Lofibra |
|  |  | Phenofibrate |
|  |  | Procetofen |
|  |  | Supralip |
|  |  | Tricheck |
|  |  | Tricor |
|  |  | Triglide |
|  | fibric acid |  |
|  | fenofibric acid | Trilipix |
|  | gemfibrozil | Lopid |

*If an individual’s EHR records indicated medications from more than one category, the largest of the corresponding constants was used to adjust median TG levels.

**Supplementary Table 24: SNPs used to build predicted gene expression for TG-associated genes, by tissue**

| **gene** | **tissue** | **rsid** | **ref_allele** | **eff_allele** |
| --- | --- | --- | --- | --- |
| *ANGPTL3* | Kidney_Cortex | rs61775882 | T | C |
|  |  | rs11207978 | T | C |
|  |  | rs72667353 | A | G |
|  |  | rs7550306 | A | C |
|  |  | rs56115463 | C | A |
|  |  | rs76150158 | T | G |
|  |  | rs61775945 | C | T |
|  |  | rs12118253 | G | A |
|  |  | rs10789121 | C | T |
|  |  | rs1168100 | T | C |
|  |  | rs912540 | G | A |
|  |  | rs10889356 | G | A |
|  |  | rs80148997 | C | T |
|  | Liver | rs12116657 | A | C |
|  |  | rs12118253 | G | A |
|  |  | rs67537755 | G | A |
|  |  | rs4587594 | G | A |
|  |  | rs10889356 | G | A |
| *ANGPTL4* | Artery_Tibial | rs55948703 | G | A |
|  |  | rs55994980 | G | A |
|  |  | rs2161568 | G | A |
|  |  | rs28377676 | G | A |
|  |  | rs74649801 | A | G |
|  |  | rs1674033 | A | G |
|  |  | rs76638576 | A | G |
|  |  | rs8106950 | C | T |
|  |  | rs6603123 | G | T |
|  |  | rs62124687 | C | T |
|  |  | rs28619893 | C | T |
|  |  | rs12980768 | C | T |
|  |  | rs4804061 | T | C |
|  |  | rs13344097 | C | T |
|  |  | rs36247 | T | C |
|  |  | rs139469000 | C | T |
|  |  | rs1808536 | G | A |
|  |  | rs62117490 | A | G |
|  |  | rs148680686 | CTTGTTTTTTTCTTT | C |
|  |  | rs11667516 | C | T |
|  |  | rs62121173 | C | T |
|  |  | rs10425933 | A | G |
|  |  | rs8102537 | A | C |
|  |  | rs8109800 | G | A |
|  | Kidney_Cortex | rs62119635 | T | G |
|  |  | rs250506 | C | A |
|  |  | rs2927710 | T | C |
|  |  | rs140695433 | G | A |
|  |  | rs2009733 | A | G |
|  |  | rs1808537 | A | G |
|  |  | rs62117477 | G | A |
|  |  | rs1808536 | G | A |
|  |  | rs2967614 | A | G |
|  |  | rs58761784 | T | C |
|  |  | rs2303180 | G | A |
|  | Heart_Atrial_Appendage | rs12974685 | C | T |
|  |  | rs7258466 | C | T |
|  |  | rs7248719 | T | G |
|  |  | rs2042899 | C | T |
|  |  | rs139469000 | C | T |
|  |  | rs1808536 | G | A |
|  |  | rs62117489 | C | A |
|  |  | rs2967614 | A | G |
|  |  | rs2913974 | A | C |
|  |  | rs2967609 | C | A |
|  |  | rs74256561 | T | C |
|  |  | rs58761784 | T | C |
|  |  | rs73000664 | G | A |
| *APOA5* | Pancreas | rs2512593 | C | T |
|  |  | rs2078684 | C | T |
|  |  | rs7949507 | G | T |
|  |  | rs7124374 | T | G |
|  |  | rs7945051 | A | C |
|  |  | rs56159851 | C | T |
|  |  | rs4938271 | T | C |
|  |  | rs508994 | C | A |
|  |  | rs17119639 | G | A |
|  |  | rs561496 | A | G |
|  |  | rs4936349 | A | G |
|  |  | rs547021 | T | C |
|  |  | rs514112 | A | G |
|  |  | rs67215893 | G | A |
|  |  | rs143291574 | C | T |
|  |  | rs7931384 | T | C |
|  |  | rs11216022 | C | T |
|  |  | rs7104866 | C | T |
|  |  | rs1240660 | A | G |
|  |  | rs112305884 | C | T |
|  |  | rs75542613 | G | A |
|  |  | rs4018880 | CAT | C |
|  |  | rs4938353 | A | G |
|  |  | rs522645 | C | A |
|  |  | rs608840 | G | A |
|  |  | rs12282721 | G | A |
|  |  | rs515756 | T | G |
|  |  | rs67023719 | C | T |
|  |  | rs681160 | T | C |
|  |  | rs11216384 | T | C |
|  |  | rs79136761 | G | A |
|  |  | rs3903091 | T | C |
|  |  | rs11603937 | G | A |
|  |  | rs75349886 | A | G |
|  |  | rs7928623 | T | C |
|  |  | rs10892147 | T | C |
| *APOC3* | Kidney_Cortex | rs11215720 | G | A |
|  |  | rs11215722 | A | C |
|  |  | rs374729 | G | A |
|  |  | rs372453 | G | A |
|  |  | rs1076141 | C | T |
|  |  | rs7927297 | G | A |
|  |  | rs548046 | C | T |
|  |  | rs76338956 | C | T |
|  |  | rs12293413 | A | G |
|  |  | rs28412270 | T | C |
|  |  | rs565622 | A | C |
|  |  | rs7110240 | T | C |
|  |  | rs17119743 | T | G |
|  |  | rs11215951 | C | T |
|  |  | rs150762661 | A | C |
|  |  | rs11215954 | G | T |
|  |  | rs925151 | G | A |
|  |  | rs58089984 | T | G |
|  |  | rs2849174 | G | A |
|  |  | rs2727788 | C | A |
|  |  | rs2854116 | T | C |
|  |  | rs2070668 | G | T |
|  |  | rs45619137 | G | A |
|  |  | rs4938369 | C | T |
|  |  | rs10790185 | C | T |
|  |  | rs1574433 | C | T |
|  |  | rs557053 | C | T |
|  |  | rs11216444 | A | G |
|  |  | rs61697909 | G | A |
|  |  | rs11216505 | G | A |
|  |  | rs76099780 | T | C |
|  |  | rs10790211 | G | A |
|  | Heart_Left_Ventricle | rs11215863 | C | A |
|  |  | rs57187567 | A | G |
|  |  | rs11216416 | C | T |
|  |  | rs1893900 | C | T |
|  |  | rs11274556 | CAGAAATCTTTTCTTAGT | C |
|  |  | rs10892154 | C | A |
| *LPL* | Kidney_Cortex | rs1388941 | G | A |
|  |  | rs1316738 | A | G |
|  |  | rs112329866 | T | G |
|  |  | rs57077102 | G | A |
|  |  | rs1837844 | C | T |
|  |  | rs28522810 | C | T |
|  |  | rs326 | A | G |
|  |  | rs28550053 | A | G |
|  | Whole_Blood | rs13276898 | C | T |
|  |  | rs6982465 | T | C |
|  |  | rs4921999 | T | C |
|  |  | rs4922000 | T | C |
|  |  | rs7839041 | A | G |
|  |  | rs4922040 | G | T |
|  |  | rs73212588 | G | A |
|  |  | rs17480272 | T | C |
|  |  | rs111690117 | A | G |
|  |  | rs111698088 | A | G |
|  |  | rs1561747 | A | C |
|  |  | rs7818070 | G | A |
|  |  | rs295 | A | C |
|  |  | rs115849089 | G | A |
|  |  | rs4128744 | C | T |
|  |  | rs76722925 | A | G |
|  |  | rs28597716 | A | G |
|  |  | rs7015766 | C | T |
|  |  | rs13273454 | C | T |
|  |  | rs10090992 | T | G |
|  |  | rs34760348 | AG | A |
|  |  | rs17092176 | T | C |
